# Identifying the Influencing Factors for Cataract Surgery Uptake in Malaysia

**DOI:** 10.1101/2025.01.29.25321323

**Authors:** Mohamad Aziz Salowi, Nyi Nyi Naing, Tay Ju Fan, Wan Radziah Wan Nawang, Siti Nurhuda Sharudin, Norasyikin Mustafa, Nor Fariza Ngah

## Abstract

**Background/Aims:** Six simultaneous population surveys using the Rapid Assessment of Avoidable Blindness (RAAB) technique, a World Health Organisation (WHO) - recommended methodology, were done in Malaysia in 2014 (National Eye Survey, NES II). The findings revealed discrepancies in the country’s key eye care regional indicators. Mobile outreach community programs were piloted in Sarawak and the Eastern Region as part of a post-survey action plan. Despite being endorsed for outreach service brands and receiving regular operational funds, hospital and community-based cataract surgery data differed between regions. We postulate that this disparity could be due to community-related factors and their interactions. Follow-up surveys were done in both regions in 2023 (NES III), and findings were compared to the previous survey. The eligible subjects in the RAAB survey were consecutively recruited for a questionnaire interview to identify factors influencing cataract surgery uptake.

**Methods:** RAAB involved a multistage cluster sampling method, with each cluster comprising 50 residents aged 50 years and older. Subjects with Pinhole Visual Acuity (PinVA) worse than 6/18, either due to cataract or following cataract surgery, were consecutively identified and interviewed using a validated questionnaire focused on Knowledge, Perception, Attitude and Practice (comprising six factors and 22 response items), along with demographic and socioeconomic variables. The data were subsequently analysed using multiple logistic regression methods.

**Results:** A total of 1,119 subjects (Eastern = 711, Sarawak = 408) were recruited. The identified factors that influenced individuals with unilateral operable cataracts to have “no surgery” in Sarawak included their “perception to own sight” [AOR: 0.67, 95% CI (0.53, 0.84) *P*=0.001] and “attitude towards treatment [AOR:1.47, 95% CI (1.17, 1.85) *P*=0.001].” Meanwhile, in the Eastern region, the factors were “perception to own sight” [AOR: 0.67, 95% CI (0.53, 0.84) *P*=0.001], “attitude towards treatment” [AOR:1.47, 95% CI (1.17, 1.85) *P*=0.001], and “practice towards information” [AOR: 1.23, 95% CI (1.01, 1.50) *P*=0.042].

For subjects with bilateral operable cataract in Sarawak, the factors that influenced them to have “no surgery” were “knowledge on surgery” [AOR: 0.35, 95% CI (0.25, 0.50) *P*<0.001], “perception to own sight” [AOR: 1.48, 95% CI (1.15, 1.89) *P*=0.002], ethnicity (Chinese compared to Malays) [AOR: 0.19, 95% CI (0.04, 0.88) *P*=0.033] and level of education (primary school compared to secondary school or above) [AOR: 5.54, 95% CI (1.49, 20.69) *P*=0.011]. Additionally, for Eastern region, the factors identified were “knowledge on surgery” [AOR: 0.35, 95% CI (0.26, 0.48) *P*<0.001] and “practice on surgery” [AOR: 0.72, 95% CI (0.62, 0.84) *P*<0.001].

**Conclusion:** The common factors influencing the cataract surgical uptake for both Sarawak and Eastern Region include “perception to own sight”, “attitude towards treatment”, and “knowledge on surgery”. Ethnicity and level of education factors were specific to Sarawak. “Practice towards information” and “practice on surgery” were specific to the Eastern Region.

## Introduction

Malaysia signed the World Health Assembly resolution (WHA66.4: Towards Universal Eye Health: A Global Action Plan 2014–2019) during the 66^th^ WHA Geneva in May 2013. By signing the documents, the country pledged commitment to implementing the items in the resolution and monitoring its progress [1,2]. One of the initiatives taken at the country level soon after adopting the resolution was to provide equal and equitable healthcare access to the population. Based on the discrepancies of key findings between the administrative regions during the National Eye Survey in 2014 (NES II) [3], the government launched *Klinik Katarak-Kementerian Kesihatan Malaysia* (Cataract Clinic Ministry of Health Malaysia *KK-KKM*) as a community cataract pilot project in Sarawak (the Malaysian Borneo) and the Eastern Region of Peninsular Malaysia aiming to address the issue of high prevalence of blindness and cataract blindness [**Fig 1**]. The objective, concept and operational strategies of the program were audited and then endorsed by the Case Study process of the Western Pacific WHO Innovation Challenge initiative in 2021/2022 [4]. Despite the establishment of service brands and the allocation of specific funds to the project, there remains a disparity in hospital and community-based data pertaining to cataract surgery (including patient profiles, surgeon practices, and key indicators for service performance and uptake) between the two regions, with the Eastern Region constantly underperforming [5–7].

**Fig 1:**
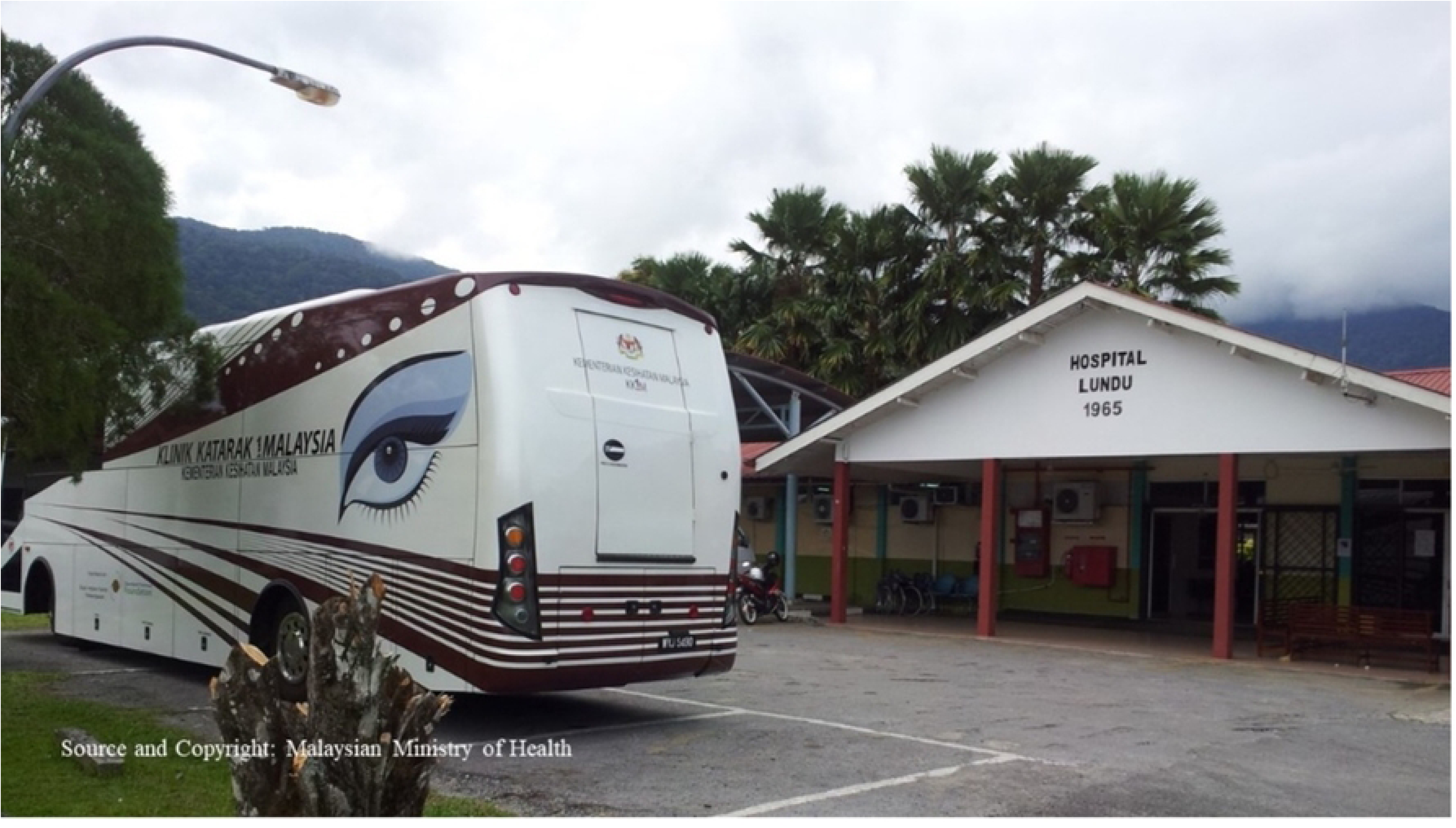
The Mobile Unit arrived at one of the locations in Sarawak.

We aim to analyse the association between the predictors and factors with the outcome (surgery done and surgery not done) in the community during the follow-up population survey after nine years of *KK-KKM* implementation in Sarawak and the Eastern Region. The findings from the study could potentially be used to formulate new policies or strengthen and optimise the implementation of the *KK-KKM* program, hence addressing the issue of service outcome discrepancy between the regions.

## Material and methods

This was a cross-sectional population survey using the Rapid Assessment of Avoidable Blindness (RAAB) technique to obtain data on eye health, specifically focusing on cataract surgery status. Additionally, structured interviews were conducted through a questionnaire to identify various factors that may influence surgical status, including knowledge, perceptions, attitudes, practices, as well as demographic and socioeconomic characteristics.

This study was divided into two phases. Phase 1 was the development and validation of a questionnaire. The factors and questionnaires were developed from an extensive literature search, expert brainstorming sessions, content validity and face validity. This process was followed by validation and reliability analysis using exploratory factor analysis (EFA), reliability tests, confirmatory factor analysis (CFA) and test-retest to produce a questionnaire named CatSurg-U Questionnaire (Cat = Cataract; Surg=Surgery; U=Uptake). It contained six factors and 22 questionnaire items [**Table 1**]. Cronbach’s alpha values of all the factors varied from 0.502 to 0.72 (total = 0.72). CFA revealed a moderate absolute and reasonable parsimonious fit model.

**Table 1:**
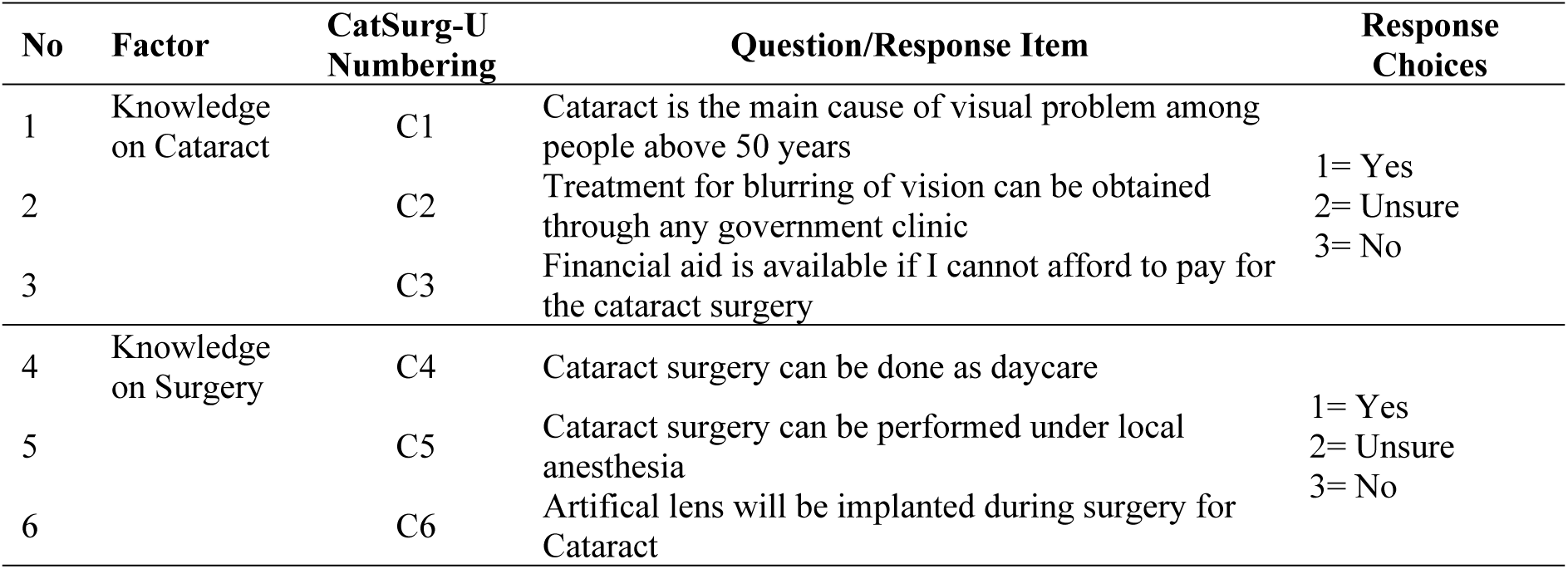
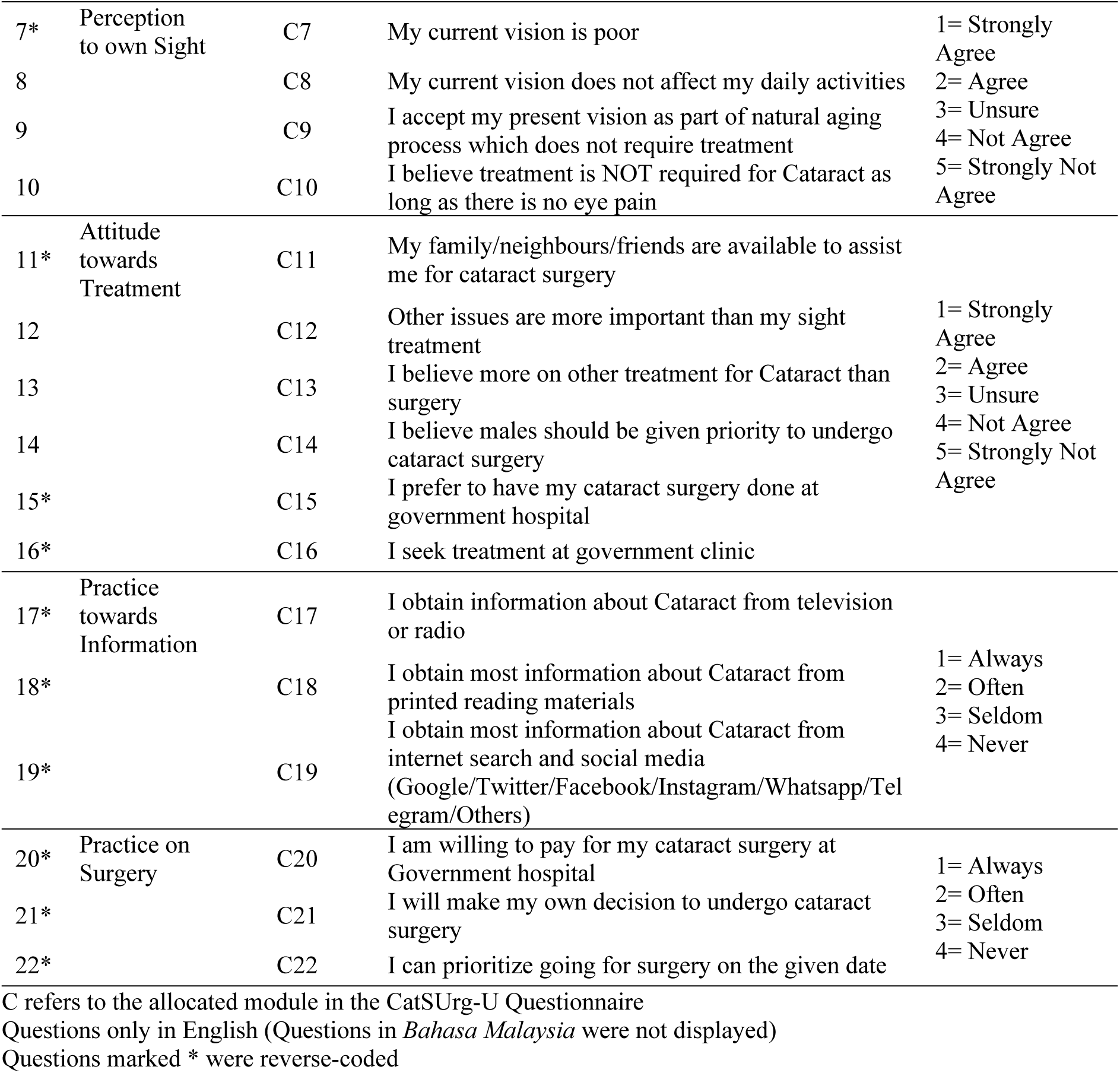
Factors and Questionnaire Items (22 items) – for CatSurg-U data collection.

Phase 1 was followed by Phase 2, a survey using the RAAB method to obtain population data on eye health and blindness [8,9]. The survey’s methodology, prevalence and cataract surgical outcome have been published [10,11]. Eligible subjects from the same sample were further interviewed using the validated CatSurg-U Questionnaire to identify factors influencing the cataract surgery uptake in both regions.

### RAAB methodology concept/summary

The Rapid Assessment of Avoidable Blindness (RAAB) is a simple and rapid cross-sectional population survey methodology that can provide investigators with data on the prevalence and causes of blindness, cataract surgical coverage and outcomes after cataract surgery. This methodology focuses on individuals aged 50 and older, thereby minimizing the sample size requirements while effectively targeting the demographic with the highest prevalence of blindness [8]. RAAB deploys a simple but focused and important eye examination technique to obtain data on the eye health of the subjects. It involves multistage cluster sampling, starting at the Department of Statistics level with a sampling frame using the concept of probability proportionate to size (PPS). However, subsequent sampling of subjects within the clusters, enumeration, examination, and interviews are done simultaneously in the field. The data analysis process is automated upon entry of information into the RAAB 7 software [12]. This system is cost-effective, as the time required to collect data within any given cluster is minimal, and it does not necessitate the use of expensive ophthalmic equipment.

Additionally, the survey can be conducted accurately and reliably by ophthalmology trainees and optometrists. The data collected from the survey serve a crucial role in designing and monitoring eye care programs within the surveyed regions [8,9].

### The sampling frame for the RAAB Survey

Department of Statistics, Malaysia (DOSM), once every ten years, conducts data collection for the National Population and Housing Census. An Enumeration Block (EB), a population unit, with 80-100 houses/residents each, is outlined according to the population distribution, based on the latest findings during the survey. This is followed by the corresponding geographical map drawing, indicating the exact location and borders of each EB. The EBs are gazetted for fieldwork activities such as morbidity, nutrition, household expenditure, and labour force surveys [13–15].

For this study, all EBs from the 2020 national census were used to select clusters for the RAAB. A total of 98 EBs for Sarawak and 105 EBs were randomly selected for the Eastern Region regardless of strata, using the Probability Proportionate to Size (PPS) technique. Individual EB codes and the corresponding maps were then brought to the field to be used to locate the EBs for data collection.

### Training for RAAB Survey and CatSurg-U Questionnaire

Each region had six (6) teams of data collectors comprising three persons: two doctors and one allied health staff member trained in ophthalmology. The training for the survey teams was a comprehensive process, conducted separately in each region one week before the fieldwork. Both sessions were led by the first author, a certified Western Pacific RAAB trainer, to monitor data quality and adherence to study protocol. The survey team members were required to attend four training days, including RAAB and CatSurg-U Questionnaire lectures, inter-observer or inter-interviewer variation, IOV (inter-interviewer reliability) assessment for both RAAB (IOV RAAB) and CatSurg-U (IOV CatSUrg-U) and a pilot survey in one of the nearby EBs during fieldwork. Each region had one coordinator responsible for the survey’s smooth implementation and progress monitoring. IOV assessments were done separately for the RAAB and CatSurg-U Questionnaire. The method and kappa results for IOV RAAB have been addressed in other publications. [10,11]

### IOV CatSurg-U

During training, the assessment for IOV CatSurg-U was conducted after the IOV RAAB on subjects, who were a mix of hospital staff and patients from the outpatient department. The subjects were different from the recruited subjects of IOV RAAB. As per the questionnaire, 22 questions were asked by six interviewers representing each survey team. Twenty-two (22) subjects in Sarawak and 20 in the Eastern Region volunteered to be interviewed. The subjects included individuals with mixed normal and impaired vision, including cataracts and pseudophakia (had been operated on for cataracts). They were interviewed by rotation (each subject was interviewed six times using the same questionnaire starting at different chronological numbers). The agreements were analysed using Fleiss Kappa [16]. For Sarawak, two items had a fair agreement, ten had a moderate agreement, eight had a substantial agreement, and two had an almost perfect agreement. For the Eastern Region, six items had a fair agreement, seven had a moderate agreement, seven had a substantial agreement, and two had an almost perfect agreement. For both regions, C8 and C9 had only fair agreement. All agreements were statistically significant [**Table 2**]. Similar to the intervention taken after IOV RAAB, post-mortem interviews and group retraining focusing on the disagreements were done before the team went to the field the next day for an actual survey. Each team member was re-explained regarding each question’s aim and trained on the best possible way to administer it to the subjects. The emphasis was also given only to allow the assigned person to conduct the interview throughout the survey.

**Table 2:**
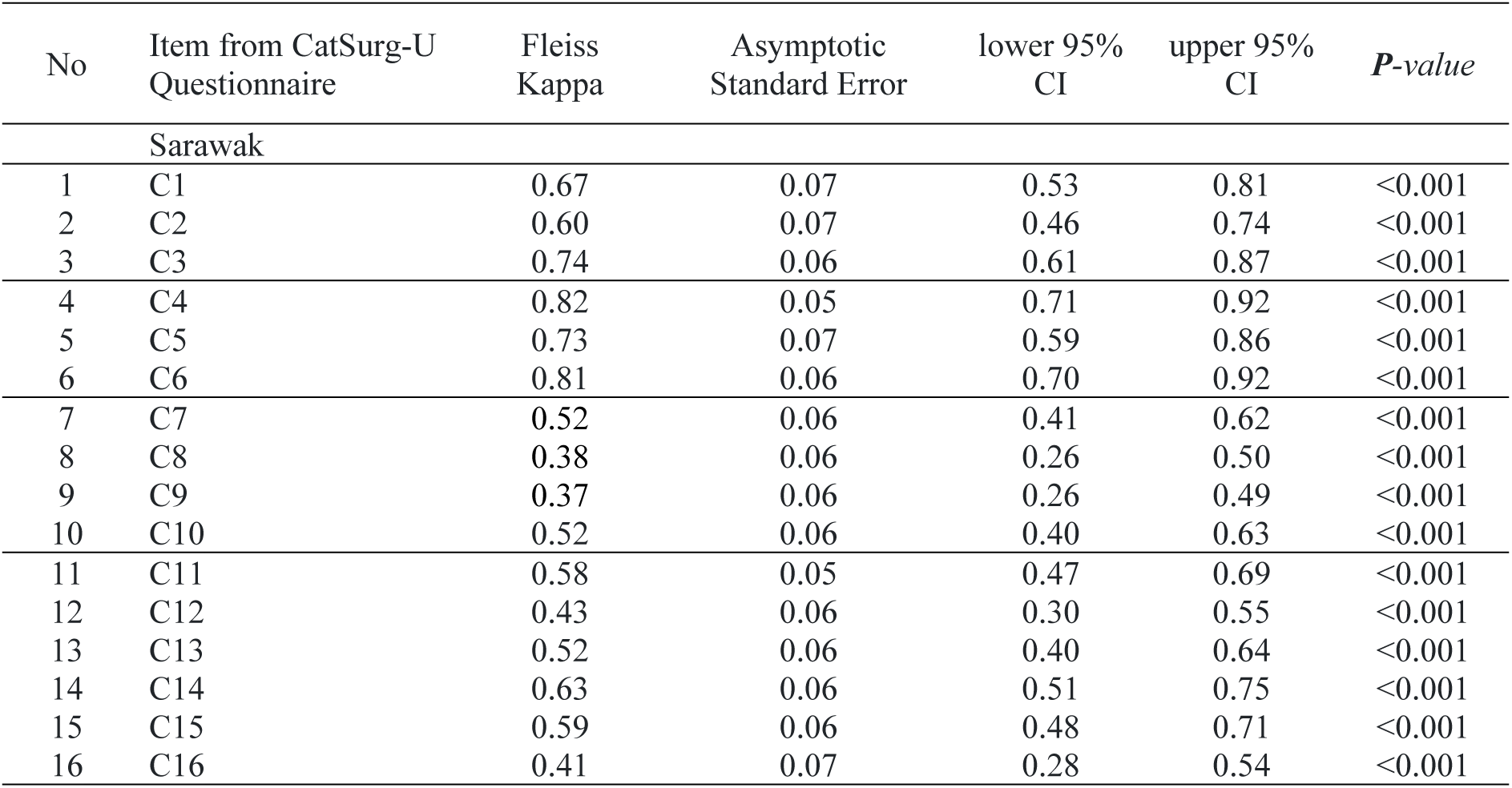
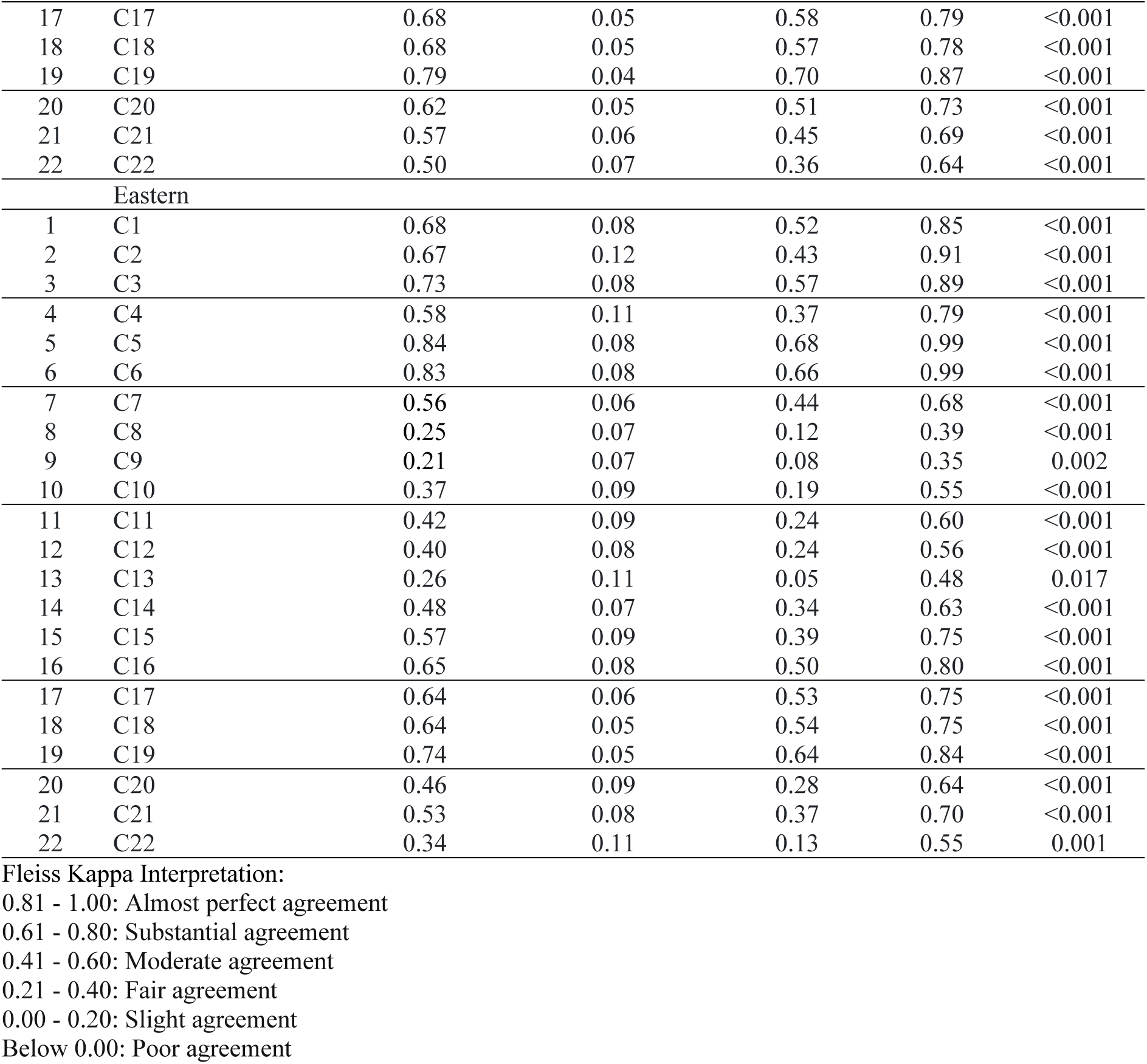
Inter-interviewer Reliability Assessment of Six Interviewers of Sarawak and the Eastern Region for agreement on CatSurg-U Questionnaire; Fleiss Kappa by Response Item (22 Questions)

| No | Item from CatSurg-U Questionnaire | Fleiss Kappa | Asymptotic Standard Error | lower 95% CI | upper 95% CI | *P-value* |
| --- | --- | --- | --- | --- | --- | --- |
| Sarawak |  |  |  |  |  |  |
| 1 | C1 | 0.67 | 0.07 | 0.53 | 0.81 | <0.001 |
| 2 | C2 | 0.60 | 0.07 | 0.46 | 0.74 | <0.001 |
| 3 | C3 | 0.74 | 0.06 | 0.61 | 0.87 | <0.001 |
| 4 | C4 | 0.82 | 0.05 | 0.71 | 0.92 | <0.001 |
| 5 | C5 | 0.73 | 0.07 | 0.59 | 0.86 | <0.001 |
| 6 | C6 | 0.81 | 0.06 | 0.70 | 0.92 | <0.001 |
| 7 | C7 | 0.52 | 0.06 | 0.41 | 0.62 | <0.001 |
| 8 | C8 | 0.38 | 0.06 | 0.26 | 0.50 | <0.001 |
| 9 | C9 | 0.37 | 0.06 | 0.26 | 0.49 | <0.001 |
| 10 | C10 | 0.52 | 0.06 | 0.40 | 0.63 | <0.001 |
| 11 | C11 | 0.58 | 0.05 | 0.47 | 0.69 | <0.001 |
| 12 | C12 | 0.43 | 0.06 | 0.30 | 0.55 | <0.001 |
| 13 | C13 | 0.52 | 0.06 | 0.40 | 0.64 | <0.001 |
| 14 | C14 | 0.63 | 0.06 | 0.51 | 0.75 | <0.001 |
| 15 | C15 | 0.59 | 0.06 | 0.48 | 0.71 | <0.001 |
| 16 | C16 | 0.41 | 0.07 | 0.28 | 0.54 | <0.001 |

| No | Item from CatSurg-U<br>Questionnaire | Fleiss<br>Kappa | Asymptotic<br>Standard Error | lower 95%<br>CI | upper 95%<br>CI | *P-value* |
| --- | --- | --- | --- | --- | --- | --- |
| 17 | C17 | 0.68 | 0.05 | 0.58 | 0.79 | <0.001 |
| 18 | C18 | 0.68 | 0.05 | 0.57 | 0.78 | <0.001 |
| 19 | C19 | 0.79 | 0.04 | 0.70 | 0.87 | <0.001 |
| 20 | C20 | 0.62 | 0.05 | 0.51 | 0.73 | <0.001 |
| 21 | C21 | 0.57 | 0.06 | 0.45 | 0.69 | <0.001 |
| 22 | C22 | 0.50 | 0.07 | 0.36 | 0.64 | <0.001 |
| Eastern |  |  |  |  |  |  |
| 1 | C1 | 0.68 | 0.08 | 0.52 | 0.85 | <0.001 |
| 2 | C2 | 0.67 | 0.12 | 0.43 | 0.91 | <0.001 |
| 3 | C3 | 0.73 | 0.08 | 0.57 | 0.89 | <0.001 |
| 4 | C4 | 0.58 | 0.11 | 0.37 | 0.79 | <0.001 |
| 5 | C5 | 0.84 | 0.08 | 0.68 | 0.99 | <0.001 |
| 6 | C6 | 0.83 | 0.08 | 0.66 | 0.99 | <0.001 |
| 7 | C7 | 0.56 | 0.06 | 0.44 | 0.68 | <0.001 |
| 8 | C8 | 0.25 | 0.07 | 0.12 | 0.39 | <0.001 |
| 9 | C9 | 0.21 | 0.07 | 0.08 | 0.35 | 0.002 |
| 10 | C10 | 0.37 | 0.09 | 0.19 | 0.55 | <0.001 |
| 11 | C11 | 0.42 | 0.09 | 0.24 | 0.60 | <0.001 |
| 12 | C12 | 0.40 | 0.08 | 0.24 | 0.56 | <0.001 |
| 13 | C13 | 0.26 | 0.11 | 0.05 | 0.48 | 0.017 |
| 14 | C14 | 0.48 | 0.07 | 0.34 | 0.63 | <0.001 |
| 15 | C15 | 0.57 | 0.09 | 0.39 | 0.75 | <0.001 |
| 16 | C16 | 0.65 | 0.08 | 0.50 | 0.80 | <0.001 |
| 17 | C17 | 0.64 | 0.06 | 0.53 | 0.75 | <0.001 |
| 18 | C18 | 0.64 | 0.05 | 0.54 | 0.75 | <0.001 |
| 19 | C19 | 0.74 | 0.05 | 0.64 | 0.84 | <0.001 |
| 20 | C20 | 0.46 | 0.09 | 0.28 | 0.64 | <0.001 |
| 21 | C21 | 0.53 | 0.08 | 0.37 | 0.70 | <0.001 |
| 22 | C22 | 0.34 | 0.11 | 0.13 | 0.55 | 0.001 |

Fleiss Kappa Interpretation:
0.81 - 1.00: Almost perfect agreement
0.61 - 0.80: Substantial agreement
0.41 - 0.60: Moderate agreement
0.21 - 0.40: Fair agreement
0.00 - 0.20: Slight agreement
Below 0.00: Poor agreement

### Survey Methods for RAAB Survey

Two separate, cross-sectional, population-based surveys, which adhered to the World Health Organization (WHO)-recommended RAAB protocol, were conducted simultaneously from 1^st^ July to 31^st^ October 2023. The surveys collected data on the prevalence of visual impairment and its causes, the prevalence of cataract, visual outcomes after cataract surgery, Cataract Surgical Coverage (CSC), effective Cataract Surgical Coverage (eCSC) and refractive error coverage (REC). Each team surveyed 16-17 randomly selected clusters and examined 50 persons aged 50 years and older. The population was sampled per RAAB methodology, the WHO-recommended method for population-based surveys [8,9]. A total of 50 subjects were recruited in each EB.

All participants provided written consent prior to their involvement in the study. They were given a consent form that explained the study’s objectives, procedures, potential risks, and their rights. The form was written in clear, understandable language, and participants had adequate time to read through it and ask any questions before consenting. For illiterate participants, verbal consent was obtained and properly documented, with a witness present, in line with the study protocol.

All recruited subjects had a brief interview, during which demographic, medical, and ocular history data were taken. It was followed by a three-meter visual acuity assessment using the RAAB7 application on tablets [12]. The doctors in the team examined the subjects’ eyes using a hand-held ophthalmoscope. If they were detected to have a visual impairment, the primary cause was identified. They were then referred to the nearest ophthalmic care facility for further management. The details of the RAAB survey protocol and methodology have been described elsewhere [8,9].

### Survey Methods for CatSurg-U Questionnaire

Subjects with Pinhole VA of <6/18 due to cataracts or those who had undergone cataract surgery were consecutively selected to be interviewed using the CatSurg-U Questionnaire. They were only included when their criteria met the above requirement, spoke and understood Malay and/or English, was a Malaysian resident of at least six months, and gave informed consent.

Some extra demographic variables were collected in addition to the variables collected for RAAB (ethnicity, marital status, occupation, education level and monthly income).

Anonymous data were recorded and uploaded for central compilation centrally using Google Forms.

### Definition of Visual Acuity

Visual Impairment (VI) categories were defined according to the Visual Acuity (VA) thresholds used in the WHO’s International Classification of Diseases (ICD-11) [17]. Blindness: VA less than 3/60 in the better eye; Severe VI: VA less than 6/60 to 3/60 in the better eye; Moderate VI: VA less than 6/18 to 6/60 in the better eye and Mild VI: VA less than 6/12 to 6/18 in the better eye. The level used to recruit subjects for the CatSurg-U Questionnaire was moderate VI or below.

### Sample size calculation

The latest population data was obtained from the Malaysian National Census 2020. [13,14] A prevalence of blindness of 1.5 % in the Eastern Region and 1.6% in Sarawak in subjects aged 50 and older from NES II (2014) was used in the calculation using a 95% confidence interval, precision of 30.0%, estimated design effect (DEFF) of 1.5 and 20.0% possibility of non-responders. [8,9] The calculation resulted in a sample size of 98 clusters (4900 subjects aged 50 years and older) and 105 clusters (5239 subjects aged 50 years and older) for Sarawak and the Eastern Region, respectively. The subjects for the CatSurg-U Questionnaire were recruited consecutively from the randomly recruited subjects in the RAAB survey. Using binary logistic regression, the sample size was calculated using Event Per Variable (EPV) as we intended to analyse the association of socioeconomic and demographic variables, factors and outcomes. EPV is defined as the ratio of the number of outcome events (no surgery) to the number of predictor variables or factors in the logistic regression model. Following the rule of thumb, 10 events per variable were required to achieve stable and reliable estimates. For example, if there were five (5) predictor variables or factors in the model, and we aimed for a minimum of 10 events per variable, the required number of events (no surgery) would be: required events = 10 ×number of predictors variables or factors, therefore required events = 10 × 5 = 50 events [18].

For this study, the required number of events (no surgery) was calculated by unilateral/bilateral eye and by the region: Unilateral (Sarawak: number of variables = 8, the required number of events = 80, Eastern: number of variables = 6, the required number of events = 60) and bilateral (Sarawak: number of variables = 12, the required number of events = 120, Eastern: number of variables = 9, the required number of events = 90). Only the required event obtained for unilateral Sarawak was below the required sample size (*n*=37). It was due to difficulties in the sampling technique and was discussed in the limitation.

### Statistical Analysis

Statistical analysis was performed using IBM SPSS AMOS Statistical Package for Social Science (SPSS), version 26.0 (SPSS, Inc., Chicago, III., USA) for Windows.

Inter-interviewer reliability assessment for the CatSurg-U Questionnaire was done using Fleiss Kappa. Binary Logistic Regression was done to identify the association of the predictor variables or factors (demographic, socioeconomic, knowledge, perception, attitude and practice factors) to the dependent outcome (surgery done vs surgery not done with the latter as the outcome event) in one or both eyes (unilateral vs bilateral) [**Fig 2**]. The analysis started with data exploration and cleaning, followed by simple logistic regression (SLogR) to screen for important independent variables. Variables with *P* value < 0.25 and/or clinically important were selected for multiple logistic regression (MLogR) using forward and backward selection techniques to produce a preliminary main effect model. The modelling procedure was done using the Likelihood Ratio test based on Maximum Likelihood Estimates (MLEs). From SLogR to MLogR, variable selection was done by four major principles, (a) best fit, (b) parsimonious model, (c) biological plausibility, and (d) statistical significance. This step was followed by a multicollinearity & interaction check to produce a preliminary final model. The Hosmer–Lemeshow goodness-of-fit test, classification tables and area under the receiving operator characteristic (ROC) curve were examined to ensure the model’s fitness before the regression model was confirmed. The results were interpreted using crude and adjusted Odds Ratios, 95% confidence intervals and corresponding p-values [19,20].

**Fig 2:**
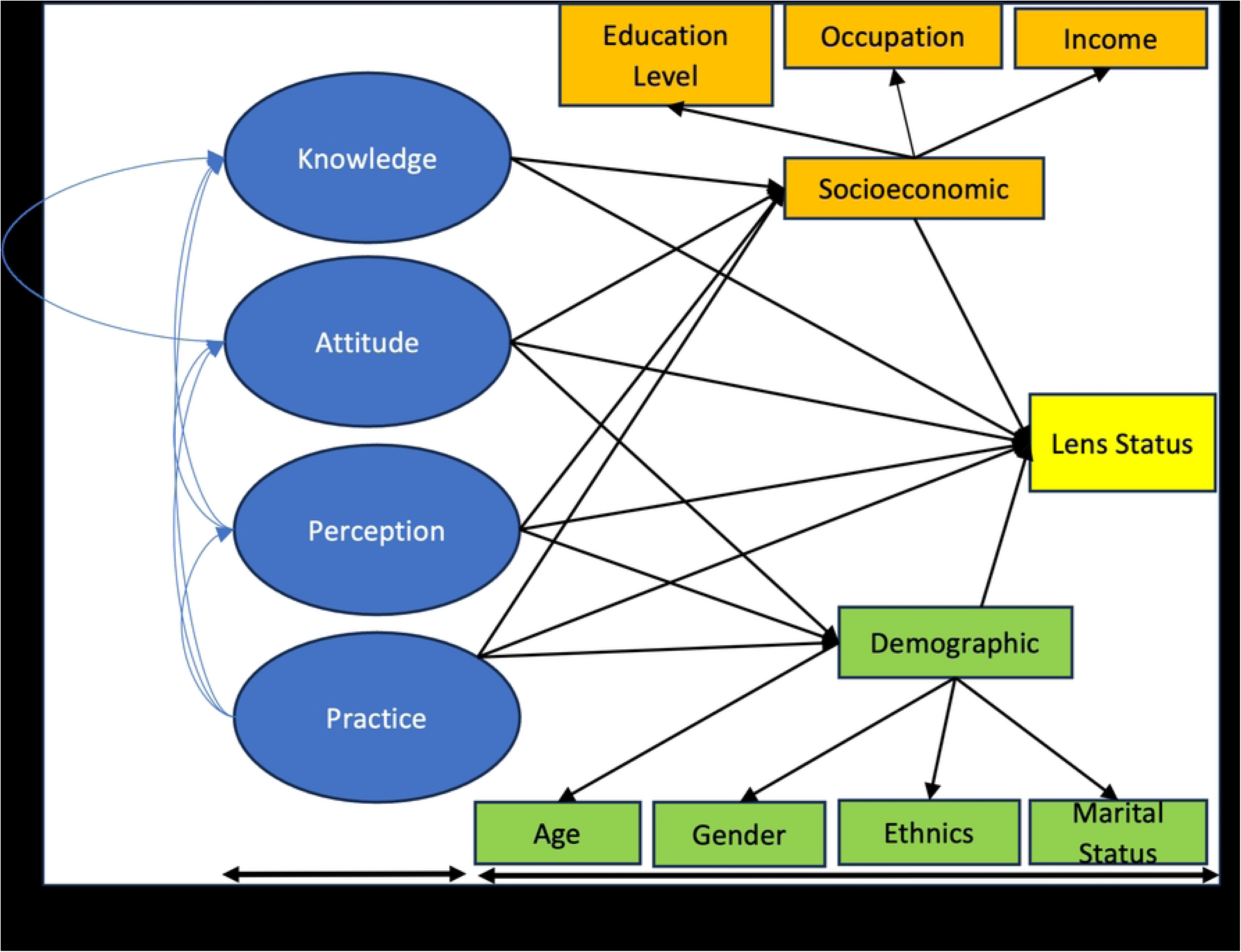
Conceptual Framework.

## Results

Some of the results related to the CatSurg-U Questionnaire development, validation and the RAAB survey are still under review or have been published. This manuscript will only address the CatSurg questionnaire data collection results following the RAAB survey. A total of 1,119 subjects (Sarawak = 408, Eastern = 711) were recruited. The demographic profile of the eligible subject for the CatSurg-U Questionnaire showed a significantly higher proportion of self-employed females, females with no formal educational background and females with income <RM1500 per month in Sarawak. There was a significantly higher proportion of Malay females and a higher significant proportion of males with primary school qualifications in the Eastern Region. There was a significantly higher proportion of married males in both regions [**Table 3**].

**Table 3:** Demographic profile of subjects recruited for the Catsurg-U Questionnaire by gender (Sarawak *n*=408, Eastern *n* = 711)

|  | SARAWAK |  |  |  |  |  |  | EASTERN |  |  |  |  |  |  |
| --- | --- | --- | --- | --- | --- | --- | --- | --- | --- | --- | --- | --- | --- | --- |
| | *N* | Female | | Male | | $\chi^2$Statistic(df) | *P*-value | *N* | Female | | Male | | $\chi^2$Statistic(df) | *P*-value |
| **Age Group:** |  |  |  |  |  |  |  |  |  |  |  |  |  |  |
| 50-59 | 32 | 15 | 6.5 | 17 | 9.7 | 4.99 (3) | 0.173 | 72 | 43 | 10.5 | 29 | 9.6 | 5.41 (3) | 0.144 |
| 60-69 | 128 | 71 | 30.6 | 57 | 32.4 |  |  | 250 | 143 | 34.9 | 107 | 35.5 |  |  |
| 70-79 | 182 | 101 | 43.5 | 81 | 46.0 |  |  | 258 | 138 | 33.7 | 120 | 39.9 |  |  |
| 80 and above | 32 | 45 | 19.4 | 21 | 11.9 |  |  | 131 | 86 | 21.0 | 45 | 15.0 |  |  |
| **Ethnicity (EASTERN):** |  |  |  |  |  |  |  |  |  |  |  |  |  |  |
| Malay |  |  |  |  |  |  |  | 623 | 368 | 90.0 | 255 | 85.0 | 7.97 (2) | 0.019 |
| Chinese |  |  |  |  |  |  |  | 65 | 27 | 7.0 | 38 | 13.0 |  |  |
| Others (including Indian, Bidayuh or Orang Ulu) |  |  |  |  |  |  |  | 23 | 15 | 4.0 | 8 | 3.0 |  |  |
| **Ethnicity (SARAWAK):** |  |  |  |  |  |  |  |  |  |  |  |  |  |  |
| Malay | 107 | 58 | 25.0 | 49 | 27.8 | 5.30 (5) | 0.380 |  |  |  |  |  |  |  |
| Chinese | 73 | 38 | 16.4 | 35 | 19.9 |  |  |  |  |  |  |  |  |  |
| Indian | 5 | 2 | 0.9 | 3 | 1.7 |  |  |  |  |  |  |  |  |  |
| Iban | 118 | 77 | 33.2 | 41 | 23.3 |  |  |  |  |  |  |  |  |  |
| Bidayuh | 34 | 19 | 8.2 | 15 | 8.5 |  |  |  |  |  |  |  |  |  |
| Others (including Orang Ulu) | 71 | 38 | 16.4 | 33 | 18.8 |  |  |  |  |  |  |  |  |  |
| **Marital Status:** |  |  |  |  |  |  |  |  |  |  |  |  |  |  |
| Married | 310 | 155 | 66.8 | 155 | 88.1 | 24.78 (1) | <0.001 | 531 | 281 | 68.5 | 250 | 83.1 | 19.35 (1) | <0.001 |
| Unmarried (including divorcee, widow or widower) | 98 | 77 | 33.2 | 21 | 11.9 |  |  | 180 | 129 | 31.5 | 51 | 16.9 |  |  |
| **Occupation:** |  |  |  |  |  |  |  |  |  |  |  |  |  |  |
| Government | 46 | 20 | 8.6 | 26 | 14.8 | 6.78 (2) | 0.034 | 39 | 19 | 4.6 | 20 | 6.6 | 5.66 (2) | 0.059 |
| Private | 18 | 7 | 3.0 | 11 | 6.3 |  |  | 42 | 18 | 4.4 | 24 | 8.0 |  |  |
| Self | 344 | 205 | 88.4 | 139 | 79.0 |  |  | 630 | 373 | 91.0 | 257 | 85.4 |  |  |
| **Level of Education:** |  |  |  |  |  |  |  |  |  |  |  |  |  |  |
| None | 163 | 115 | 49.6 | 48 | 27.3 | 35.73 (3) | <0.001 | 210 | 156 | 38.0 | 54 | 17.9 | 40.58 (3) | <0.001 |
| | *N* | Female | | Male | | $\chi^2$Statistic(df) | *P*-value | *N* | Female | | Male | | $\chi^2$Statistic(df) | *P*-value |
| Primary School | 146 | 83 | 35.8 | 63 | 35.8 |  |  | 290 | 153 | 37.3 | 137 | 45.5 |  |  |
| Secondary School | 74 | 29 | 12.5 | 45 | 25.6 |  |  | 177 | 91 | 22.2 | 86 | 28.6 |  |  |
| Diploma or above | 25 | 5 | 2.2 | 20 | 11.4 |  |  | 34 | 10 | 2.4 | 24 | 8.0 |  |  |
| **Average Household Monthly Income:** |  |  |  |  |  |  |  |  |  |  |  |  |  |  |
| RM1500 or less | 196 | 121 | 52.2 | 75 | 42.6 | 8.16 (3) | 0.043 | 281 | 163 | 39.8 | 118 | 39.2 | 3.51 (3) | 0.319 |
| >RM1500-RM3000 | 129 | 75 | 32.3 | 54 | 30.7 |  |  | 257 | 157 | 38.3 | 100 | 33.2 |  |  |
| >RM3000-RM6000 | 56 | 24 | 10.3 | 32 | 18.2 |  |  | 135 | 70 | 17.1 | 65 | 21.6 |  |  |
| >RM6000 | 27 | 12 | 5.2 | 15 | 8.5 |  |  | 38 | 20 | 4.4 | 18 | 5.3 |  |  |

*df*=Degree of freedom
All were tested using the Chi-Square test.

There were two common significant factors identified which influenced the individuals with unilateral operable cataract to have “no surgery” in both regions (“perception to own sight” and “attitude towards treatment”). The other significant factor was “practice towards information” (specific to the Eastern Region) [**Table 4**].

**Table 4:**
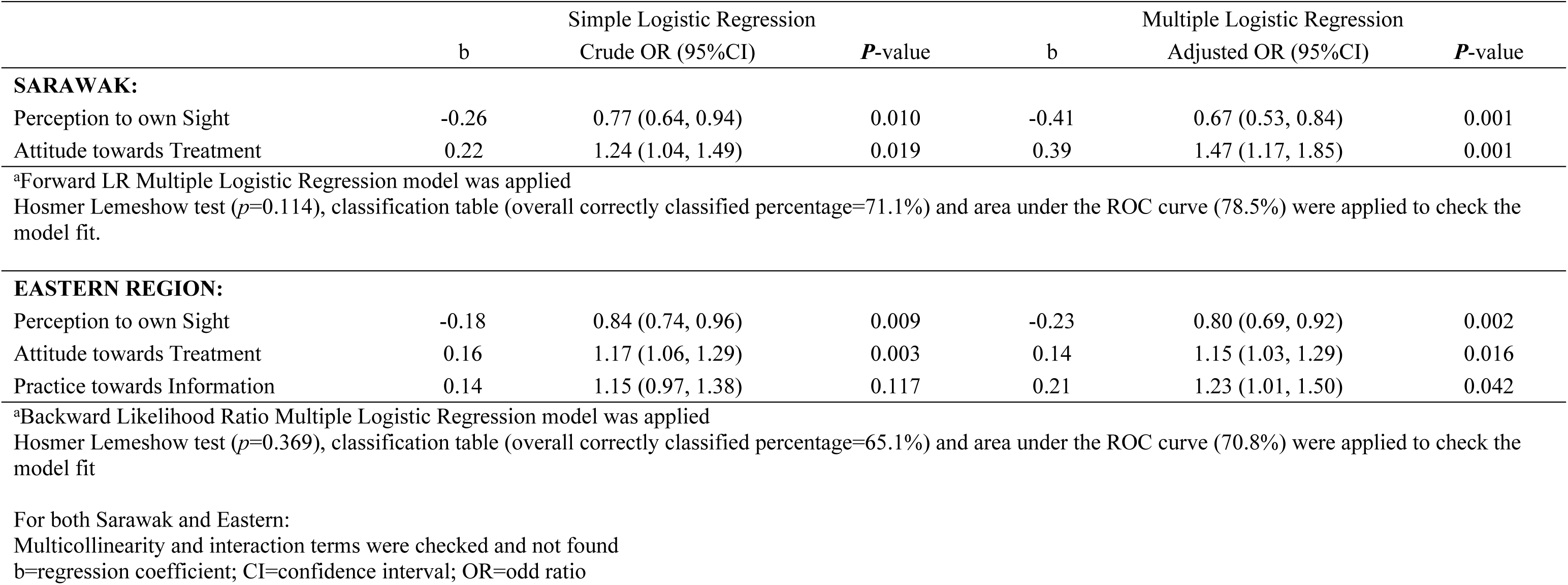
Associated Factors Influencing Individuals with UNILATERAL (one eye) Operable Cataract to have “No Surgery” by Simple and Multiple Logistic Regression Model (Sarawak *n*=90, Eastern *n*=212)

In Sarawak:

1. Individuals with better “perception to own sight” had 33.0% lower odds of having no surgery when they had an operable cataract in one eye [AOR: 0.67, 95% CI (0.53, 0.84) *P*=0.001, adjusted for “attitude towards treatment”].
2. Individuals with better “attitude towards treatment” had 47.0% higher odds of having no surgery when they had an operable cataract in one eye [AOR:1.47, 95% CI (1.17, 1.85) *P*=0.001, adjusted for “perception to own sight”].

In the Eastern Region:

1. Individuals with better “perception to own sight” had 20.0% lower odds of having no surgery when they had an operable cataract in one eye [AOR: 0.80, 95% CI (0.69, 0.92) *P*=0.002, adjusted for “attitude towards treatment” and “practice towards information”].
2. Individuals with better “attitude towards treatment” had 15.0% higher odds of having no surgery when they had an operable cataract in one eye [AOR: 1.15, 95% CI (1.03, 1.29) *P=*0.016, adjusted for “perception to own sight” and “practice towards information”].
3. Individuals with better “practice towards information” had 23.0% higher odds of having no surgery when they had an operable cataract in one eye [AOR: 1.23, 95% CI (1.01, 1.50) *P*=0.042, adjusted for “perception to own sight” and “attitude towards treatment”].

For bilateral operable cataract, the common significant factor that influenced the individuals to have “no surgery” in both regions was “knowledge on surgery”. The other significant factors were “perception to own sight”, ethnicity and level of education (specific to Sarawak) and “practice on surgery” (specific to the Eastern Region) [**Table 5**].

**Table 5:** Associated Factors Influencing Individuals with BILATERAL (both eyes) Operable Cataract to have “No Surgery” by Simple and Multiple Logistic Regression Model (Sarawak *n*=243, Eastern *n*=352)

|  | b | Simple Logistic Regression<br>Crude OR (95%CI) | P-value | b | Multiple Logistic Regression<br>Adjusted OR (95%CI) | P-value |
| --- | --- | --- | --- | --- | --- | --- |
| **SARAWAK:** |  |  |  |  |  |  |
| **Ethnicity:** |  |  |  |  |  |  |
| Malay | 0 | 1 | <0.001 | 0 | 1 | 0.016 |
| Chinese | -1.72 | 0.18 (0.07, 0.48) | 0.001 | -1.67 | 0.19 (0.04, 0.88) | 0.033 |
| Iban | 1.02 | 2.76 (0.79, 9.64) | 0.111 | 1.48 | 4.41 (0.76, 25.73) | 0.100 |
| Others (including Bidayuh, Indian or Orang Ulu) | -0.02 | 0.98 (0.34, 2.08) | 0.973 | 0.41 | 1.50 (0.37, 6.04) | 0.567 |
| **Level of Education:** |  |  |  |  |  |  |
| None | 2.50 | 12.13 (4.62, 31.83) | <0.001 | 0.31 | 1.36 (0.32, 5.71) | 0.677 |
| Primary School | 2.11 | 8.26 (3.35, 20.38) | <0.001 | 1.71 | 5.54 (1.49, 20.69) | 0.011 |
| Secondary School or above | 0 | 1 |  | 0 | 1 | 0.028 |
| Knowledge on Surgery | -0.95 | 0.39 (0.30, 0.50) | <0.001 | -1.05 | 0.35 (0.25, 0.50) | <0.001 |
| Perception to own Sight | -0.32 | 1.38 (1.16, 1.63) | <0.001 | 0.39 | 1.48 (1.15, 1.89) | 0.002 |
| Hosmer Lemeshow test ($p=0.480$), classification table (overall correctly classified percentage=93.0%) and area under the ROC curve (93.2%) was applied to check the model fit | | | | | | |
| **EASTERN REGION:** |  |  |  |  |  |  |
| Knowledge on Surgery | -1.27 | 0.28 (0.21, 0.38) | <0.001 | -1.05 | 0.35 (0.26, 0.48) | <0.001 |
| Practice on Surgery | -0.57 | 0.56 (0.50, 0.64) | <0.001 | -0.33 | 0.72 (0.62, 0.84) | <0.001 |

Hosmer Lemeshow test ( $p=0.126$ ), classification table (overall correctly classified percentage=84.4%) and area under the ROC curve (90.4%) were applied to check the model fit
For both Sarawak and Eastern:
\*Forward Likelihood Ratio Multiple Logistic Regression model was applied
Multicollinearity and interaction terms were checked and not found
b=regression coefficient; CI=confidence interval; OR=odd ratio

In Sarawak:

1. Individuals with better “knowledge on surgery” had 65.0% lower odds of having no surgery when they had operable cataract in both eyes [AOR: 0.35, 95% CI (0.25, 0.50) *P*<0.001, adjusted for ethnicity, level of education and “perception to own sight”].
2. Individuals with better “perception to own sight” had 48.0% higher odds of having no surgery when they had operable cataract in both eyes [AOR: 1.48, 95% CI (1.15, 1.89) *P*=0.002, adjusted for ethnicity, level of education and “knowledge of surgery”].
3. Chinese individuals had 81.0% lower odds, compared to Malay individuals, of having no surgery when they had operable cataract in both eyes [AOR: 0.19, 95% CI (0.04, 0.88) *P*=0.033, adjusted for level of education, “knowledge of surgery” and “perception to own sight”].
4. Individuals who attended primary school had 5.5 times higher odds, compared to those who attended secondary school or above, of having no surgery when they had operable cataract in both eyes [AOR: 5.54, 95% CI (1.49, 20.69) *P*=0.011, adjusted for ethnicity, “knowledge of surgery” and “perception to own sight”].

In the Eastern Region:

1. Individuals with better “knowledge on surgery” had 65.0% lower odds of having no surgery when they had operable cataract in both eyes [AOR: 0.35, 95% CI (0.26, 0.48) *P*<0.001, adjusted for “practice of surgery”].
2. Individuals with better “practice on surgery” had 28.0% lower odds of having no surgery when they had operable cataract in both eyes [AOR: 0.72, 95% CI (0.62, 0.84) *P*<0.001, adjusted for “knowledge of surgery”].

## Discussion

The KK-KKM project in both regions emphasises scheduled trips for screening and surgery and revisiting them after one month by optometrists to assess patients’ visual outcomes. The timetable for the mobile unit is distributed to all the Provincial Hospitals at the beginning of each calendar year. The fixed schedule allows people in remote areas to plan their finances and trips to come forward and seek eye treatment. Operating in proper operating rooms using standard cataract extraction techniques, adhering to the quality measurement of biometry and fixing the timetable for the service maximise access and ensure quality surgery for the people.

Like all other hospital facilities in the Malaysian Ministry of Health, data from cataract surgeries performed at the *KK-KKM* locations are entered into the National Eye Database, a web-based password-protected surveillance system collecting data on eye diseases and the clinical performance of the ophthalmology program in Malaysia. It consists of online systematic data entry according to predefined sets of preoperative, operative and outcome forms. Details on the Malaysian Cataract Surgery Registry and Cumulative Summation (CUSUM) Techniques in cataract surgical performance monitoring have been published elsewhere [21,22]. The main reason behind these strategic measures was to ensure maximum access to quality cataract surgeries for every individual in the region regardless of differences in socioeconomic and demographic profiles.

This study combined the RAAB survey findings (surgery status obtained from ocular examination by trained eyecare providers) with the CatSurg-U Questionnaire findings (factors and response/questionnaire items), demographic and socioeconomic data [Fig 2]. Peng et al. (2013), in their population survey to evaluate the utilisation of eye care services in a rural population in North China, used the frequency of visits to eye care providers as the outcome of multiple logistic regression in analysing the factors associated with the underuse of services [23]. Li et al. (2020), in a cross-sectional survey in rural Yueqing, Wenzhou, China, used a questionnaire on knowledge, attitudes and practices (KAP). Visual acuity (unilateral vs bilateral) was used as the outcome variable of multiple logistic regression to identify factors affecting the use of eye care services [24]. To the best of our knowledge, no available study in the literature yet evaluates the uptake of cataract surgery using an objective outcome measurement (surgery status) obtained from ocular examination.

The following were the outcome variables collected during the fieldwork (RAAB survey) obtained from the ocular examination. They were evaluated and discussed with the research committee before the commencement of the survey:

I. Lens status
variable was obtained from direct eye examination using a handheld direct ophthalmoscope. The findings included normal, opacity, aphakia, pseudophakia with no posterior capsular opacity, and pseudophakia with posterior capsular opacity or no view. Cataract surgery uptake was represented by either aphakia, pseudophakia (surgery done), or opacity (surgery not done).
II. Distant pinhole visual acuity (PinVA)
variable was obtained by testing the subjects’ distant visual acuity with pinholes using the built-in chart in the RAAB7 apps. The findings were an option of the levels of PinVA (6/12, 6/18, 6/60, 3/60. 1/60, Perception of light or No perception of light). The inclusion criteria for the CatSurg Questionnaire interview was Pinhole VA worse than 6/18.
III. Cataract surgery

Four sub-variables represented this variable:

a. Age when the surgery was done (numerical)
b. Type of cataract surgery [categorical: options were no Intraocular Lens (IOL), IOL implantation or couching]
c. Cost of cataract surgery (categorical: options were totally free, partially free or fully paid)
d. Cause of poor vision after cataract surgery (categorical: options were ocular co-morbidity, operative complication, refractive error or long-term complications)

Unlike Variable II and III, Variable I had a clear binary definition between “surgery done” and “surgery not done”. For Variable II, subjects could have undergone surgery, but the visual acuity remained poor (e.g. surgery with intra-operative or post-operative complication); likewise, they could have lens opacity, but the visual acuity remained good. Variable III only captured subjects who had undergone cataract surgery (and excluded subjects who had cataracts but did not undergo surgery).

The results from the multiple regression analysis suggested that there were two common significant factors identified which influenced the individuals with unilateral operable cataract to have “no surgery” in both regions (“perception to own sight” and “attitude towards treatment”). The other significant factor was “practice towards information” (specific to the Eastern Region). For bilateral operable cataract, the common significant factor that influenced the individuals to have “no surgery” in both regions was “knowledge on surgery”. The other significant factors were “perception to own sight,” ethnicity, and level of education (specific to Sarawak), as well as “practice on surgery” (specific to the Eastern Region).

In Sarawak, having better “perception to own sight” was associated with lower odds of “no surgery” in unilateral cataract. However, the factor was associated with higher odds if the person had a bilateral cataract. This contradictory finding in the “perception to own sight” between unilateral and bilateral operable cataract could be explained by the awareness of the individuals’ visual capacity by comparing their eyes to their fellow eyes. They would seek treatment because the visual discrepancy between the eyes could be disabling. However, when both eyes progressively developed impairment, the individual would adapt to their surroundings and falsely perceive that there was no need to seek treatment, especially when the need for good vision for daily activities in remote areas was minimal. In Sarawak, where geographical and financial access could be an issue for the community, those with bilateral operable cataract should not be neglected and must be reached out by expanding the coverage of the mobile unit.

In both regions, when a person had unilateral operable cataract, although a better “perception of own sight” was associated with lower odds of “no surgery”, the opposite was revealed for the “attitude towards treatment”. Even if the community had a positive attitude towards seeking cataract treatment, the odds of having “no surgery” were higher. They had good self-awareness about poor vision in one eye and possibly wanted to seek treatment, but access to treatment (geographical or financial) could be the limiting factor. Interpreting the “attitude towards treatment” and “practice towards information” together, within the scope of the questionnaire items (the better the attitude and practice, the higher the odds of “no surgery) in the Eastern Region pointed more to possible limited “supply” of service despite higher “demand”. A concerted effort to increase the coverage and capacity of services must be addressed to increase the uptake of cataract surgery in the population, especially in the Eastern Region.

Ethnic (Chinese, compared to Malays) and level of education (primary school, compared to secondary school or above) contributed to the factors influencing individuals in Sarawak with bilateral operable cataract to have “no surgery”. These factors were not identified in the Eastern Region. Sarawak is a multicultural state. The Iban are the largest indigenous group in Sarawak, comprising approximately 30.0-35.0% of the population, Chinese 24.0-26.0%, Malays 20.0-25.0%, Bidayuh 6.0-7.0% and other indigenous groups, including the Orang Ulu, Melanau, Kayan, Kenyah, Kelabit, and Penan, who together make up around 10.0-15.0% of the population [25]. The lower odds among Chinese with bilateral operable cataract with “no surgery” perhaps was contributed by the ethnic composition in the state and their better access to treatment (geographically). Analysis of other specific ethnics was not significant. The individuals with primary school qualifications had 5.5 times higher odds of “no surgery " than those with higher qualifications. These findings emphasised the importance of education in providing eyecare in Sarawak and other regions.

A better “knowledge on surgery” if the individuals had bilateral operable cataract was associated with lower odds of “no surgery” in both regions. This indicated that if the community had substantial knowledge about cataract surgery, the odds of them coming forward to seek treatment would be higher. Similarly, “practice on surgery” was associated with lower odds of “no surgery” when the individuals had bilateral operable cataract but only specific to the Eastern Region.

The strategies proposed to increase the uptake of cataract surgery in the community shall be targeted at the identified influencing factors associated with higher odds of “no surgery” in specific regions. In Sarawak (“attitude towards treatment” in unilateral operable cataract) and (level of education and “perception to own sight” in bilateral operable cataract). In the Eastern Region (“attitude towards treatment” and “practice towards information” in unilateral operable cataract)

Peng et al. (2013), in their evaluation of the use of eye care services in a rural population, revealed ocular comorbidities (glaucoma, age macular degeneration or refractive error) as the factors influencing the community to visit an eye care provider [23]. Li et al. (2020) stated “no need” and “schedule conflicts” as the main reasons for not seeking eye care. High education levels, older age, self-perceived vision condition and regular vision check behaviour were related to seeking eye care services [24].

The remedial measures for this study shall include expanding and upgrading cataract surgical services in both regions. This expansion/upgrade must include the hospital infrastructure, the mobile unit, and workforce capacity, especially the surgeons’ training. This expansion will allow active case detection, especially for individuals residing in remote areas with bilateral operable cataracts. Other remedial measures include strengthening community advocacy, education and engagement to empower and sustain the long-term project, especially in the Eastern Region.

### Limitation

The primary population survey in this study was NES III using the Rapid Assessment of Avoidable Blindness (RAAB) methodology. It collected eye health data (including lens or surgery status). Based on this survey, the sample size was calculated, and grants were allocated. Data collectors extracted the eligible subjects from each randomly selected Enumeration Block (EB) in NES III to be interviewed using the CatSurg-U Questionnaire. The number of subjects, therefore, depended on the availability of eligible subjects within the selected EB. Although calculated, recruiting subjects to satisfy the sample size for the intended regression analysis was difficult as the population was not sampled for the questionnaire.

### Conclusion

Intervention is required to expand and optimise the capacity of the eye care services to improve the uptake of cataract surgery, especially in the Eastern Region. A concerted effort must be taken with relevant stakeholders to educate and increase public knowledge about cataract surgery. Further research among the eyecare providers is also required to identify the latent factors affecting the country’s delivery of cataract surgery services.

## Data Availability

All study data are available from the database URL https://www.raab.world/survey-data

## Acknowledgement

The authors would like to thank the Director General of the Ministry of Health Malaysia for his permission to publish this article. Additionally, the authors acknowledge the significant contributions of data collectors involved in data entry for both NES II and NES III.

## Ethics approvals

Ethical approvals were obtained from Medical Research and Ethics Committees (MREC) of the Ministry of Health, Malaysia (Research ID NMRR-19-197-46172). The study was conducted in accordance with the tenets of the Declaration of Helsinki.

